# A prospective two-year longitudinal follow-up study depicting humoral and cell-mediated immune responses in Covaxin vaccinated individuals

**DOI:** 10.1101/2023.06.02.23290825

**Authors:** Archana Tripathy, Sreeparna Podder, Swatishree Sradhanjali, Debdutta Bhattacharya, Sanghamitra Pati, Sunil K. Raghav

**Author notes:** Corresponding author Correspondence: Sunil K. Raghav, Ph.D.

## Abstract

There are majorly two variants of SARS-CoV-2 vaccine that were employed worldwide on emergency basis to contain the COVID-19 pandemic i.e., RNA based or adenovirus construct based Spike protein expression system which was broadly used and the inactivated virus particle composition. Due to emergency usage starting from the onset of 2021, the immunogenicity data pertaining to long term effects of these vaccines is unexplored. Therefore, in this study we assessed the immunogenicity analysis of Covaxin (BBV152), an inactivated virus-based vaccine for a longitudinal time-span of two years. We investigated the humoral and cell-mediated immune responses in 250 subjects for two years by estimating the RBD specific IgG titres and CD4+/CD8+ T-cell responses. We found that anti-RBD IgG titres that were almost reaching at the basal levels within a year of 2nd dose of vaccination, went significantly high immediately after Omicron infection wave in January 2022. Moreover, the pseudo-virus neutralization by the serum of these subjects showed concordant and drastic increase in virus neutralization activity. At the same time, mild or no symptoms were observed in individuals infected with Omicron variant of SARS-CoV-2. These observations strongly suggested that Omicron variant could have been the best SARS-CoV-2 variant for effective vaccine formulations to generate robust protective immune response along with lesser side effects. Interestingly, the CD4+ and CD8+T-cell activity in Covaxin vaccinees depicted mild to moderate but sustained responses. The spike peptivator pool activated PBMCs of vaccinees depicted an enhancement of CD4+ and CD8+ antigenic responses after 2^nd^ and 3^rd^ dose of vaccine administration. In comparison to Covishield, the antibody and T-cell responses were found to be milder in BBV152 vaccinees. This milder antibody and T-cell response could be the reason behind no or less side effects with BBV152 administration than other RNA based vaccines. Overall, our study is one of the first studies profiling the longitudinal humoral and T-cell responses of inactivated virus-based vaccines like COVAXIN, which was predominantly used in India and neighbouring Southeast Asian countries.

## Introduction

The Corona virus disease-19 (COVID-19) outbreak, first reported in Wuhan, China in late December, 2019 had affected millions of people and caused significant deaths worldwide [1]. COVID-19 is a highly contagious disease caused by Severe Acute Respiratory Syndrome Coronavirus 2 (SARS-CoV-2), an enveloped RNA virus that belongs to the Coronaviridae family [2]. The World Health Organization (WHO) declared the novel coronavirus outbreak as global pandemic in March, 2020 due to its high transmission efficiency. It is transmitted through the respiratory aerosols and direct or indirect contact with the infectious particles/droplets. It elicits mild to severe respiratory illness, characterized by dry cough, cold, runny nose, sore throat, fever or chills, chest congestion, shortness of breath or pneumonia (in more severe cases) [3].

The quick genetic evolution and emergence of new variants of concern (VOC) of SARS-CoV-2 such as B.1.1.7 (Alpha), B.1.351 (Beta), P1 (Gamma), B.1.617.2 (Delta) and B.1.1.529 (Omicron), posed a considerable challenge to restrain COVID-19 due their rapid transmissibility, virulence, immune evading/escaping trait and diminished response to vaccines and therapeutics [4, 5]. Several vaccines against SARS-CoV-2 have been developed or are in the process of development around the world over the past 2-3 years [5–7]. There are currently three types of vaccines available against the SARS-CoV-2 virus with different mechanism of action namely; mRNA, adenovirus vector, and inactivated virus vaccines. Several vaccine efficacy studies have shown promising results ranging from 50%-96% effectiveness against symptomatic COVID-19 infected individuals [8–11]. However, rapid decline in vaccine efficacy from 1 to 6 months has been noticed in fully vaccinated individuals [10]. Further, the rapid emergence and spread of SARS-CoV-2 VOCs compromises the efficacy of vaccines due to their acquired potential to evade the neutralising antibodies or cell-mediated immune response. Although rare but adverse effects following vaccine administration have also been reported earlier. Consequently, detailed insight into the underlying molecular mechanism of SARS-CoV-2 infection and host immune response is of paramount importance for efficient vaccine development, taking into account, the safety of the vaccine. Several longitudinal studies of humoral antibody response induced by COVID 19 vaccines have been reported in the recent past [12–14]. However, we are the first to show the longitudinal IgG antibody profile and neutralizing antibodies (Nab) in the serum of Covaxin (BBV152) vaccinated individuals, vaccine which was predominantly given in India and neighbouring Southeast Asian countries.

In the present prospective cohort study, we deciphered the humoral immune response against SARS-CoV-2 in Covaxin vaccinated and/or convalescent individuals through longitudinal IgG antibody profiling against receptor binding domain (RBD) of ancestral strain of SARS-CoV-2. Further, the neutralizing capabilities of serum isolated from Omicron infected individuals were evaluated against the ancestral strain. Additionally, the T-cell mediated immune response (CD4+ and CD8+T cells) was also evaluated in vaccinated and/or convalescent patients against ancestral strain and its cross reactivity with Delta and Omicron variant was determined. The observations from our results suggested that the Omicron variant of SARS-CoV-2 could be a better vaccine candidate for future innovations in this aspect.

## Materials and Methods

### Study design

The longitudinal cohort study was conducted from January, 2021 to January 2023 with 250 participants from Institute of Life Sciences, Bhubaneswar, to investigate the RBD IgG antibody titre and T-cell responses against SARS-CoV-2 at seven time points (before 2^nd^ dose: T_0_, after 10 days of 2^nd^ dose: T_1_, after 3 months of 2^nd^ dose: T_2_, after 6 months of 2^nd^ dose: T_3_, after 12 months of 2^nd^ dose: T_4_, after 10 days of 3^rd^ dose: T_5_ and after 24 months of 2^nd^ dose: T_6_). We followed these enrolled participants (n=34) up to the specified timeline in order to collect blood samples and the necessary information at each time point. All participants were interviewed and their demographic information, SARS-CoV-2 infection history, and vaccination details were recorded. The study was reviewed and approved by the Institutional Ethics Committee (BT/CS0053/05/21).

### Isolation, Preservation of PBMCs and Serum sample

Human peripheral blood was collected in EDTA-coated and spray-coated silica vacutainers (BD) separately for PBMCs and serum separation under sterile condition. PBMCs were isolated from the blood samples using Ficoll-Hypaque density gradient solution (Lymphoprep, STEMCELL TECHNOLOGIES) using the manufacturer’s protocol and cryopreserved successfully under recommended conditions. The serum was aliquoted and stored in -80℃ until further use.

### Chemiluminescent Microparticle Immunoassay

Serum samples were used for the evaluation of IgG antibody against RBD region of Spike antigen by using ARCHITECT i1000SR (Abbott Diagnostics) which is based on chemiluminescent microparticle immunoassay (CLIA). Total IgG was estimated using ARCH SARS-CoV-2 IgG II Quant (Abbott Diagnostics) kit as per the manufacturer’s instruction. The cut-off value for this quantitative kit was 50 AU/ml.

### Neutralization Assay

Serum samples of vaccinated individuals at two time points (T_2_ and T_4_) were considered for neutralization assay according to manufacturer’s protocol (SARS-CoV-2 Surrogate Virus Neutralization Test Kit, Genscript). Briefly, positive control, negative control and serum samples (serum 1:2 dilution) were mixed with HRP-RBD with a volume ratio of 1:1 and incubated for 30 mins at 37℃. Sample mixture of volume 100 µl were then added to the pre-coated hACE2 plate and incubated for 15 mins at 37℃ followed by washing the wells for 4 times with 1X washing buffer. After complete removal of residual wash buffer, 100 µl of TMB solution was added to each well and incubated in dark for 15 mins at RT. Finally, 50 µl of stop solution was added to each well and absorbance was measured immediately at 450 nm using Multiskan reader (Thermo Scientific). The formula for calculating the percent inhibition/neutralization was = (1 − OD value of sample/OD value of negative control) × 100%.

### PBMC Stimulation *in Vitro*

Cryopreserved PBMCs were quickly thawed at 37℃ and resuspended in RPMI media (Gibco) supplemented with 1% Penicillin/Streptomycin solution (Gibco) and 5% heat-inactivated human AB serum (Sigma-Aldrich). Cells after washing were cultured at 1.5 x10^6^ cells per well for 12 hours in 96 well U-bottom plate. Next, PBMCs were stimulated with peptide pools of SARS-CoV-2 Wild type, Delta and Omicron (Milteyni Biotec) at 1 µg/mL for 24 hours. For, positive control cells were treated with 1 µg/mL of Cytostim (Milteyni Biotec) with same condition (Supplementary Figure 3) and unstimulated condition was considered for negative control. After, 24 hours of stimulation, cells were washed with 1X PBS and proceeded for antibody staining for FACS. Brefeldin A (10µg/ml, eBiosciences) was added to the media after 10 hours of PBMC stimulation with peptide pools for detection of intra-cellular proteins.

### Flow cytometry

The expression of cytokines and activation induced markers (AIM) by stimulated PBMCs was analysed using full spectra flow cytometer (CYTEK AURORA). Briefly, the cells were resuspended in 100µl of FACS buffer (3% FBS in PBS) and stained for viability as well as surface markers for 30 minutes in dark at 4℃ with the antibodies: anti-CD3-BV570 (BioLegend), anti-CD8-cFluor™ B532, anti-CD4-cFluor™ YG584, anti-CD45RA-cFluor™ V450 (Cytek), anti-CCR7-BV421(BioLegend). For intracellular staining, cells were washed with FACS buffer and subsequently fixed with BD Cytofix (BD Biosciences) for 20 minutes at 4℃. After washing with BD Cytoperm (BD Biosciences), cells were labelled with anti-Granzyme B-PE Cy7, anti-IFN-γ-BV605 (BioLegend), anti-CD137-BUV615, anti-CD154-BUV737 and anti-TNF-α-BUV 395 (BD Biosciences). The expression levels were measured and data was analyzed using FCS express 7 and FlowJo V10. Fluorescence minus one (FMO) control were used to determine the appropriate gates. Representative flow cytometry plots displaying the gating strategy used to assess CD4+ and CD8+ T cell subtypes in response to peptide pools is shown in Supplementary Figure 1.

### Data Analysis

Statistical analyses were performed using Graph Pad Prism 8. Significant differences were tested by unpaired Student’s t-test and one way ANOVA. The data were expressed as the mean ± SEM. A two-tailed *p* value of <0.05 was considered statistically significant. The Loess regression method was used to conduct the correlation assessment.

The percentage of AIM+ cells after stimulation with peptide pools was divided by the percentage of AIM+ cells generated from the unstimulated group to compute the stimulation index (SI). The SI <1 was represented as 1. The median plus standard deviation of the lowest observed values, like in the case of stimulation with Spike peptide pool, was used to calculate the upper limit of the positive stimulation index.

## Results

### Longitudinal RBD specific IgG profile in the Covaxin (BBV152) vaccinated subjects

To understand the changes in RBD specific IgG antibody titres after vaccination, we established a cohort of 250 subjects who were vaccinated with the Covaxin (BBV152). The antibody titres were estimated at regular 3 months intervals after 1^st^ (D1) and 2^nd^ (D2) dose administration. The serum samples were subjected to CLIA based IgG antibody estimation against wild type (WT) SARS-CoV-2 spike protein and its persistence at seven longitudinal time points were analysed; (T_0_; 28 days post 1^st^ dose (n=184), T_1;_ 10 days post 2^nd^ dose (n=123), T_2_; 3 months post 2^nd^ dose (n=92), T_3;_ 6 months post 2^nd^ dose (n=87), T_4;_ 12 months post 2^nd^ dose (n=76), T_5;_ 18 months post 2^nd^ dose & 10 days post 3^rd^ dose (n=51) and T_6;_ 24 months post 2^nd^ dose & 6 months post 3^rd^ dose (n=46). We also segregated the subjects based on their infection status before and after vaccination to assess how infection along with vaccination impacts the IgG titres. We found that irrespective of the time points till T_3_, 15-20% of subjects were non-responders with no antibody response after vaccination (Table 1). At T_0_, more than 50% subjects were non-responders and the responders also had lesser IgG titres than after 2^nd^ dose administration i.e. T_1_ (Table 1). The IgG titres were found to be significantly decreased gradually after T_1_ till T_4_, the time of Omicron variant infection wave of SARS-CoV-2 (Figure 1). Interestingly, we found that majority of the subjects at T_4_, i.e., after Omicron wave of infection depicted very high IgG titres and there were no or only few non-responders (Figure 1 A-C). The results indicated that Omicron variant has much wider antigenic epitopes on spike protein leading to higher IgG titre antibody generation, which is quite evident from the drastically enhanced number of mutations found in Omicron Spike RNA sequence as compared to WT SARS-CoV-2 or the Delta variant (Supplementary Figure 1).

**Figure 1:**
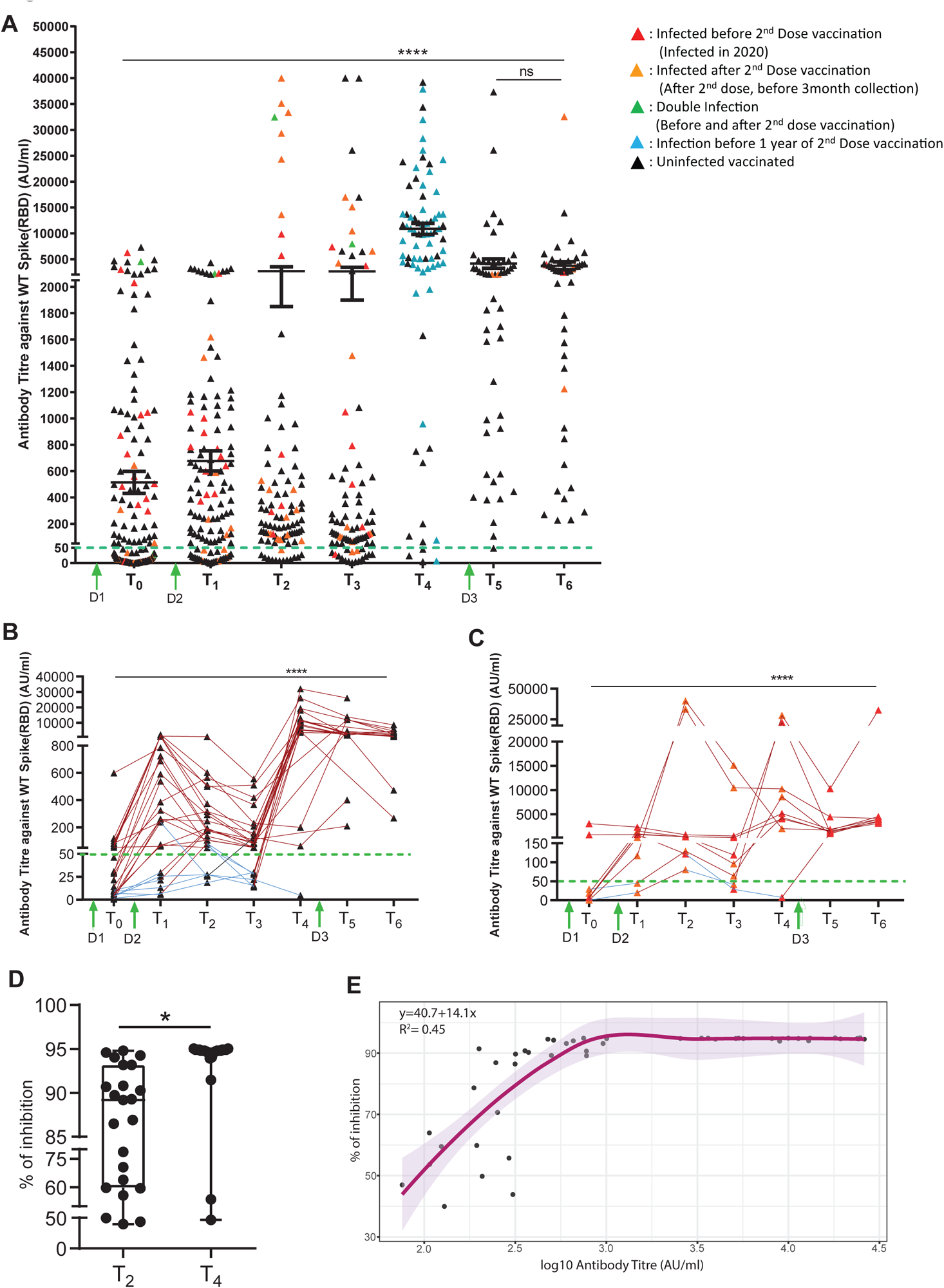
Longitudinal profiling of RBD-specific IgG antibody. (A)Vaccine induced IgG production in the vaccinated population at seven time points evaluated by performing CLIA (n=250). RBD specific IgG antibody production over the course of the trial in uninfected (B) and convalescent vaccinated (C) follow-up subjects (n=9: convalescent, n=25: uninfected). (D) Inhibition of SARS-CoV RBD–hACE2 interaction by sera from uninfected follow up individuals at 3 months and 12 months of 2^nd^dose vaccination. (E) Corelation study between antibody titre vs % of inhibition of neutralising antibodies in vaccinated individuals without any breakthrough infection by using Loess regression analysis. On the graph, each individual in the data is represented by a point and dotted lines indicating the cut-off value. Seronegative (blue lines) are regarded to be below the cut off, whereas seropositive (red lines) are considered to be over the cut off. Bars represent mean ± SEM. \**p*<0.05, \*\*\*\**p*<0.0001. Significant differences were tested by unpaired Student’s t-test and one way ANOVA with Tukey’s multiple comparison test.

**Table 1:**
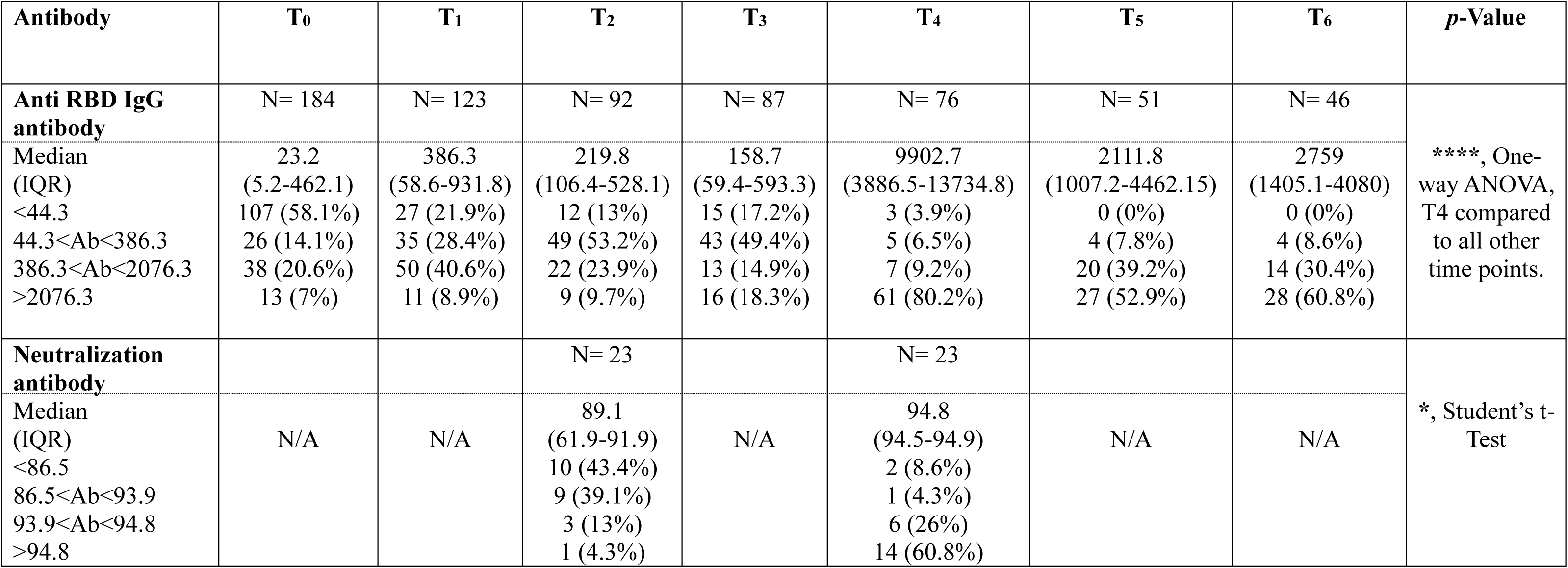
Descriptive analysis of IgG-RBD antibody titre and neutralisation antibody titre of Covaxin (BBV152) vaccinated individuals.

After T_4,_ 3^rd^ dose of vaccine was administered in June 2022, and we found that even after 3^rd^ dose of vaccination the IgG titres were significantly lesser at T_5_ than T_4_, the levels after Omicron (Figure 1). The similar level of IgG titres was the maintained till T_6._ These profiles strongly suggested Omicron variant has higher potential of generating sustained IgG antibody titres than SARS-CoV-2 variant originally used for making vaccine BBV152. Moreover, we separately looked into the IgG titre longitudinal profiles in uninfected (n=25) and convalescent individuals (n=9) based on prior infection status and we found that though infected individuals showed higher IgG titres upon Covaxin administration and the antibody titres were sustained for longer time as compared to uninfected subjects from the cohort that were vaccinated only. Interestingly both these groups behave similarly with highest antibody titres at T_4_ i.e., Omicron infection wave time (Figure 1B & 1C).

### Pseudo-virus neutralization assay to assess the virus neutralizing activity of serum from Covaxin vaccinees

It is very important to check that the higher antibody titres are corroborating with neutralization of the virus or not. Therefore, we performed spike RBD-ACE2 interaction-based surrogate virus neutralisation assay on representative samples from two interesting time points T_2_ and T_4_. As the Covaxin was generated using the WT variant of SARS-CoV-2 virus strain so we expected more neutralisation of spike-RBD by the serum of T_2_ (before Delta variant wave) as compared to T_4_, the serum of subjects after Omicron infection. We wanted to assess the serum samples’ ability to effectively neutralise COVID-19 virus. Therefore, we selected 23 subjects with follow-up sample of both time-points. We found that drastically increased antibody titres upon Omicron infection also correlated significantly and strongly in virus neutralising capacity as compared to T_4_ serum samples (Figure 1D). Subjects depicted > 95% inhibition at T_4,_ wheras the value of neutralisation capacity varied from 50% to 95% for T2 serum samples (Figure 1D). Additionally, to this significant difference, the neutralizing antibody presented a higher Interquartile Range (IQR) of 94.5-94.9 at T4, compared to T2 which had a lower IQR of 61.9-91.9. This sheds light on how Omicron presence facilitates an increased level of neutralizing antibodies against WT-RBD. Further, we performed the correlation study between antibody titre *versus* % of inhibition in vaccinated subjects without any breakthrough infection (Figure 1 E) and we observed a significant correlation of 0.45.

### Covaxin induced mild to moderate CD4+T cell responses against WT SARS-CoV-2

To test for the generation of CD4+T cell responses against WT SARS-CoV-2 following Covaxin vaccination, we considered 121 donors from the cohort. Blood samples were collected at five different time points, T_2_; 3 months, T_3_; 6 months, T_4_; 12 months, T_5_; 18 months after 2^nd^ dose & 10 days post 3^rd^ dose and T_6_; 24 months post 2^nd^dose and 6 months after 3^rd^ dose of vaccination. For this assay, the PBMCs were challenged with WT spike synthetic peptides/peptivator pool for 24h followed by FACS staining for estimation of T-cell activation and cytokine status.

After 24h of antigen challenge, the percentage of AIM+ (CD154+CD137+), AIM+EM+ (CD45RA-CCR7-), CD4+TNF-α+ and CD4+IFN-γ+ T-cells were estimated by full-spectral flow cytometry analysis. The results are represented as fold increase in the percentage positive cells as compared to unstimulated conditions (Figure 2 and Supplementary figure 2). Our data revealed that subjects vaccinated with the 2^nd^dose (T_2_) and 3^rd^dose (T_5_) of vaccine depicted enhanced AIM+ and TNF-α+ CD4 T-cells (Figure 2A and Figure 2C). Moreover, we found that TNF-α expressing CD4+T-cells were found to be significantly increased (*p* < 0.01) at T_5_ i.e., after 3^rd^ dose of vaccination as compared to T_3_, T_4_ months and T_6_ (Figure 2C). We did not find any significant change in the effector memory CD4+T-cells between different time points but the trend showed more EM cells at T_2_ and T_3_ as compared to later time points (Figure 2B). Similarly, the IFNγ+ CD4+T-cells also did not show any difference at any time-points (Figure 2D).

**Figure 2:**
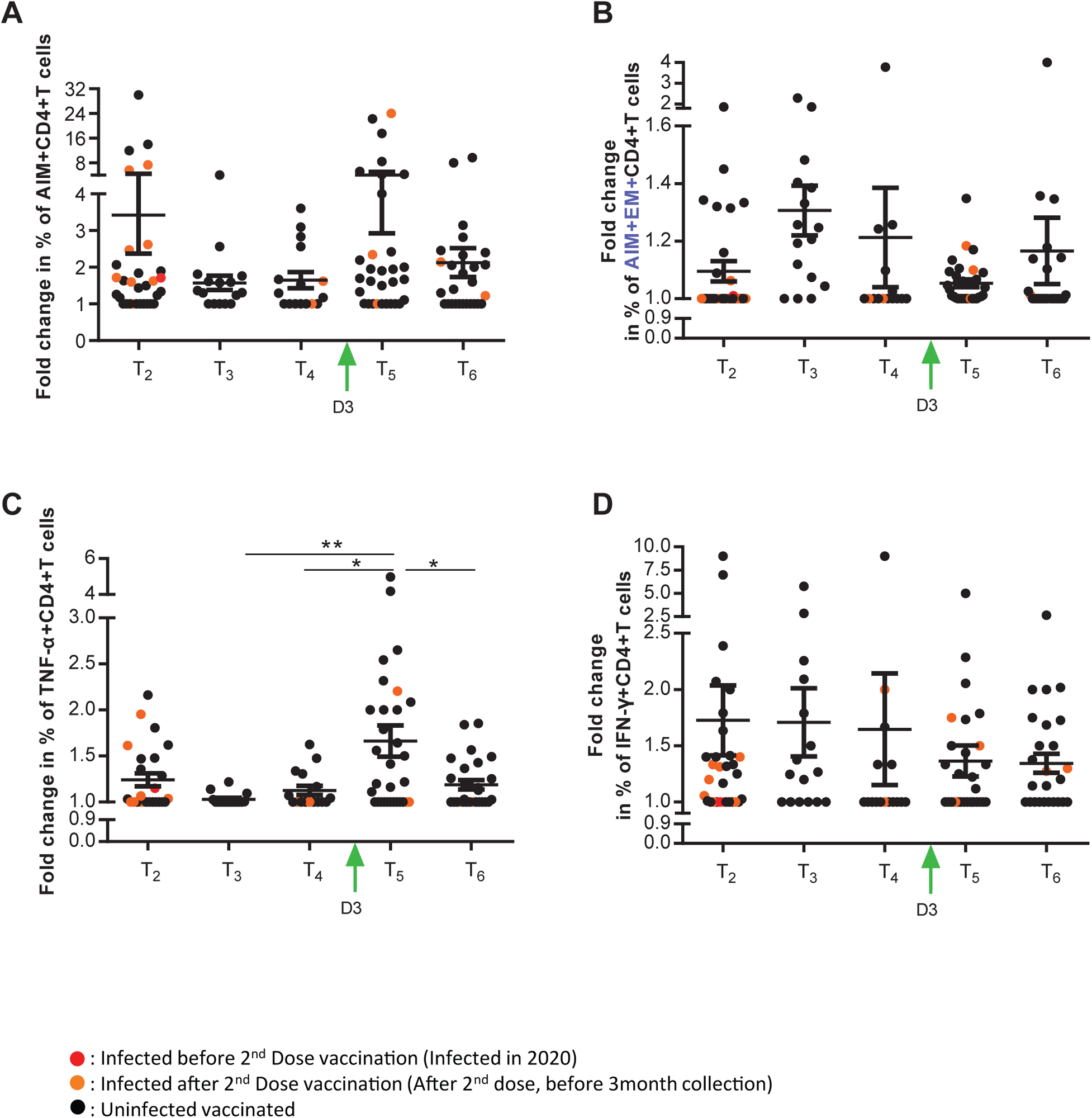
CD4+T cell responses to SARS-CoV-2 (WT) spike peptides measured by flow cytometry. The fold increase in the percentage of (A) AIM+CD4+, (B) AIM+EM+, (C) TNF-α+CD4+ and (D) IFN-γ+CD4+ have been plotted after stimulation with WT spike synthetic peptides/peptivator pool for 24 hours followed by antibody staining and flow cytometric evaluation. The graphs represent the fold increase in the percentage positive cells after normalization with their respective unstimulated controls. Data are expressed as geometric mean and SEM. Three months (n=31), six months (n=16), twelve months (n=16), eighteen months (n=31) and twenty-four months (n=27). \**p*<0.05, \*\**p*<0.01. Significant differences were tested by one way ANOVA with Tukey’s multiple comparison test.

### Delta and Omicron variants of SARS-CoV-2 depicted lesser CD4-T cell activation in Covaxin vaccinated subjects

The functionality of the vaccine induced CD4+T cells remains poorly characterized against different SARS-CoV-2 variants. Therefore, we were interested to confirm whether T-cells of Covaxin vaccinated subjects showed similar activation status with Delta and Omicron variant spike peptide pools as compared to WT SARS-CoV-2 peptide pool. To analyse CD4+T cells responses against SARS-CoV-2 major variants prevalent in India, PBMCs of 23 vaccinated individuals were stimulated with peptivator pool specific to spike of WT, Delta and Omicron.

We observed that in WT variant, the fold increase in percentage of AIM+ CD4+T-cells were much higher than in Delta and Omicron variant suggesting that Delta and Omicron variant spike peptides were varied from the WT SARS-CoV-2 and therefore resulted into increased immune-escape and infectivity (Figure 3A). Similarly, the EM+ CD4+T cells were also found to be in higher frequencies in WT variant as compared to Delta and Omicron. Though, we did not observe any noticeable change in the TNF-α, and IFN-γ producing CD4+T-cells in between variants (Figure 3C and 3D).

**Figure 3:**
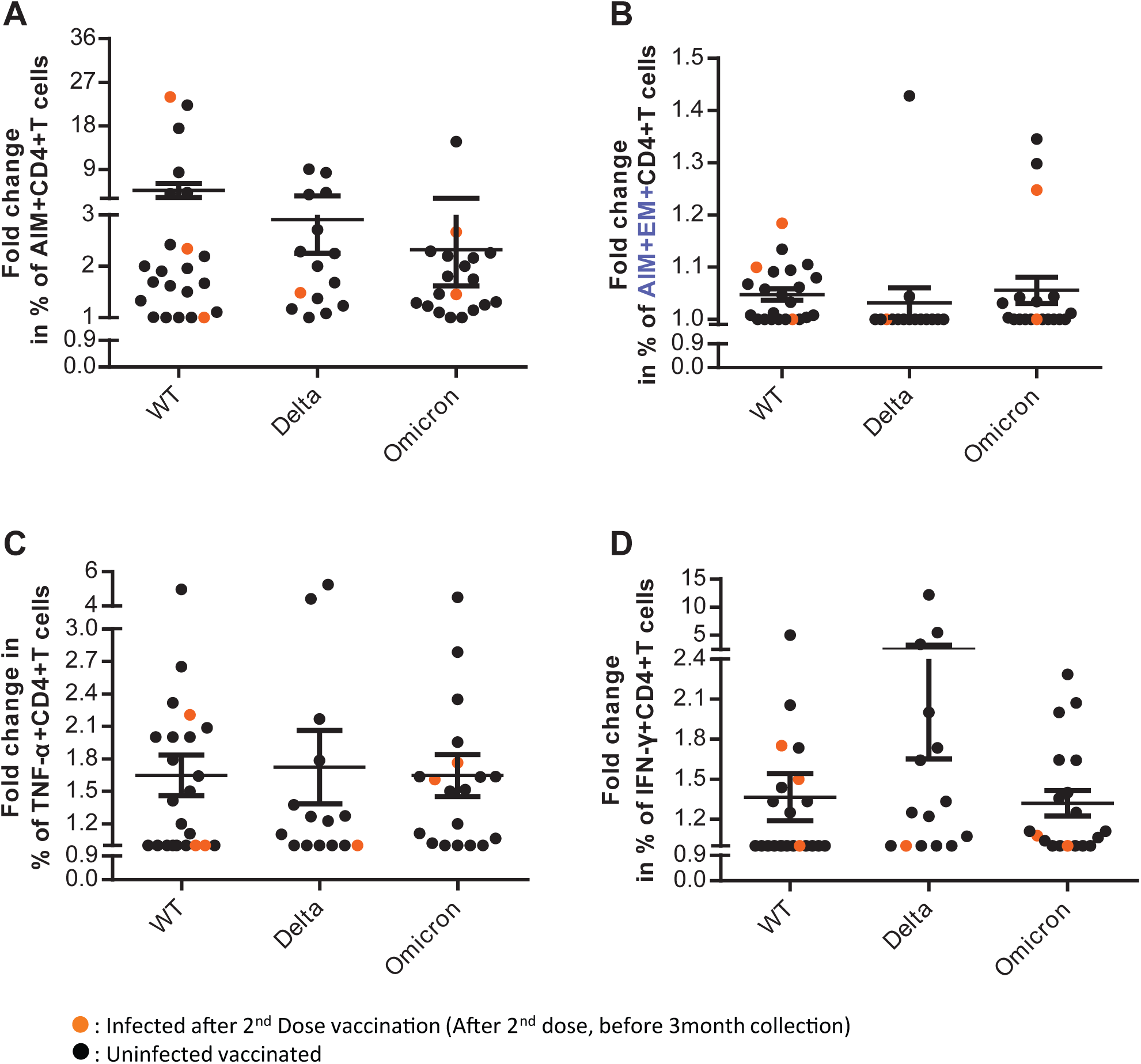
CD4+T cell responses against ancestral and mutated variants of SARS-CoV-2 spike (S) following 3^rd^ dose vaccination. PBMCs of vaccinated individuals (n=23) were stimulated with S peptivator pool corresponding to WT, Delta and Omicron. Fold increase represented in percentage of (A) AIM+ (B) AIM+EM+, (C) TNF-α+ and (D) IFN-γ+ cells. Data represented as mean ± SEM. n= 23 for spike WT, n= 15 for delta, n= 19 for omicron.

### Covaxin induced mild to moderate CD8+T cell responses against WT SARS-CoV-2

Next, we tested Cytotoxic T-cell immunity in Covaxin vaccinated individuals at five different time points, T_2_; 3 months, T_3_; 6 months, T_4_; 12 months, T_5_; 18 months after 2^nd^ dose & 10 days post 3^rd^ dose and T_6_; 24 months post 2^nd^dose and 6 months after 3^rd^ dose of vaccination. The PBMCs of 121 individuals were stimulated with WT spike peptide pool for 24 hours and stained with specific marker antibodies followed by full spectra FACS analysis. Flow cytometric evaluation was done to determine CD8+AIM+ (CD137+), AIM+TEMRA+ (CD45RA+CCR7-), AIM+EM+ (CD45RA-CCR7-), CD8+ IFN-γ+ and CD8+ Granzyme B+ T-cells (Figure 4).

**Figure 4:**
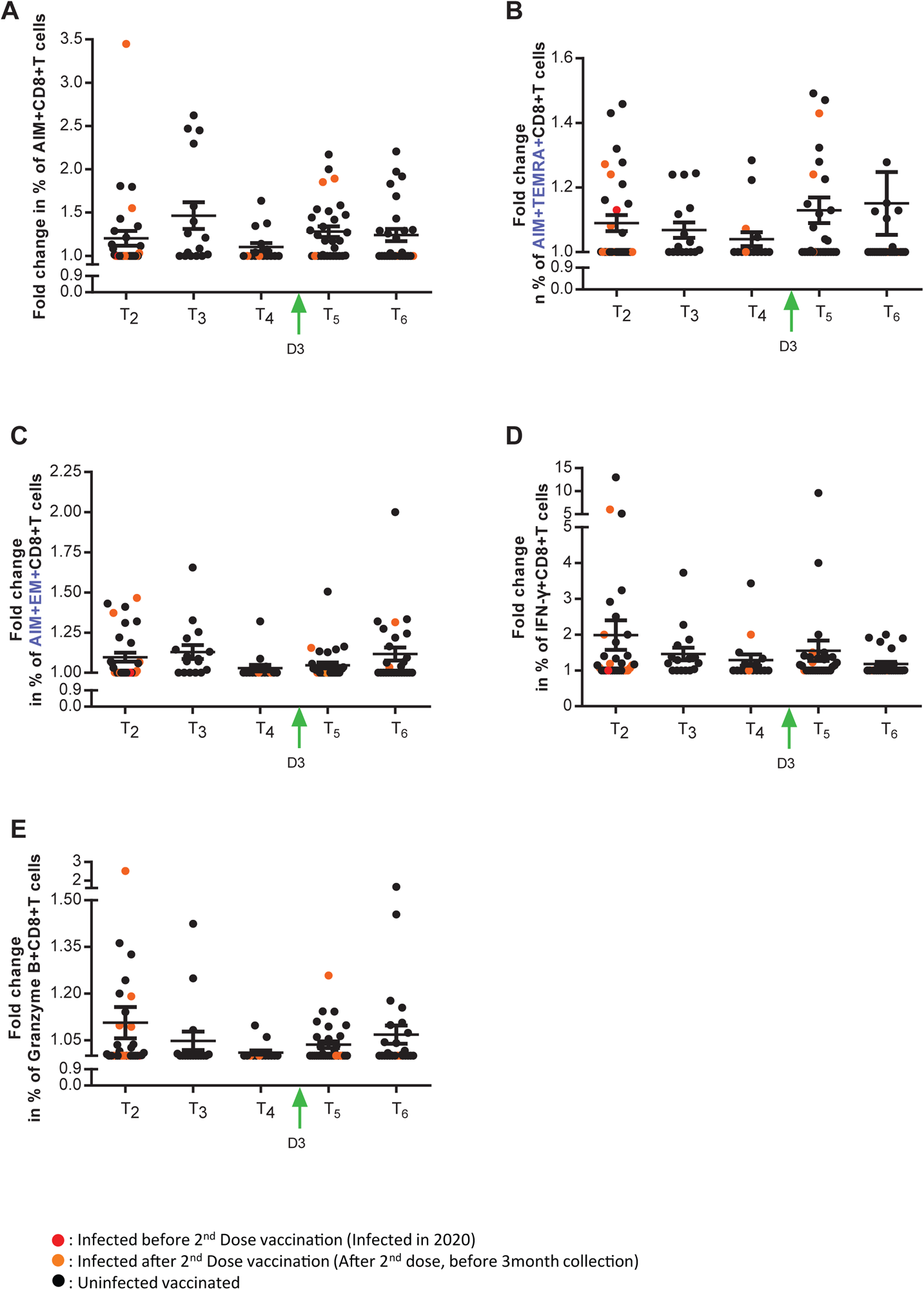
Cytotoxic T cell responses of COVID-19 vaccinees against ancestral variant (WT) Flow cytometric evaluation was done to determine fold increase in the percentage of (A) AIM+CD8+, (B) AIM+TEMRA+, (C) AIM+EM+, (D) IFN-γ+CD8+ and (E) Granzyme B+ CD8+T cells after stimulation with WT spike synthetic peptides. Data represented as mean ± SEM. Three months (n=31), six months (n=16), twelve months (n=16), eighteen months (n=31) and twenty-four months (n=27).

The results indicated that the fold-increase in the percentage of AIM+ CD8+T-cells were increased during early T_2_, T_3_ time points after vaccination, which gradually went down at T_4_ i.e. 12 months after 2^nd^ dose (Figure 4 A). Moreover, upon 3^rd^ dose of vaccination, we observed an insignificant increase in the fold-change of CD137+ CD8+T-cells at T5 and T6. On the other hand, the AIM+TEMRA+ cells showed increase after 3 months of 2^nd^ dose of vaccine (T_2_), which went down gradually till one year and then upon 3^rd^ dose these AIM+TEMRA+ cells were replenished back immediately within 10 days and then decreased back again in 6 months. The AIM+ EM+ cells, IFNγ+ and Granzyme+ CD8+T-cells also showed similar responses. Overall, the data showed a clear mild to moderate response of cytotoxic T-cells upon Covaxin administration. We did not find a statistically significant difference in cell frequencies across time points. Overall, the data revealed that the vaccination is ineffective in generating stronger CD8+T-cell immunity for a prolonged period of time.

### Different SARS-CoV2 variants fail to activate cytotoxic T cells in the vaccinated cohort

SARS-CoV-2 mutations have led to the discovery of its different variants. To analyse the effect of these variants on CD8+T cells, we stimulated PBMCs with spike peptide pool from WT, Delta and Omicron variants of SARS-CoV-2 virus for 24 hours. In this experiment, we considered 23 subjects who already received their 3^rd^dose of vaccine.

The effect of multiple SARS-CoV-2 variants upon the immunological responses for AIM+ (Figure 5 A), AIM+TEMRA+ (Figure 5 B), AIM+EM+ (Figure 5 C), IFN-γ+ (Figure 5 D) and Granzyme B+ (Figure 5 E) in CD8+ T cells were observed. The AIM+ population was detectable in 50 % individuals against WT (Figure 5 A), although it was not reflected in other variants. Overall, the results showed that vaccination increased cytotoxic response against WT while having a modest impact on Delta and Omicron variants.

**Figure 5:**
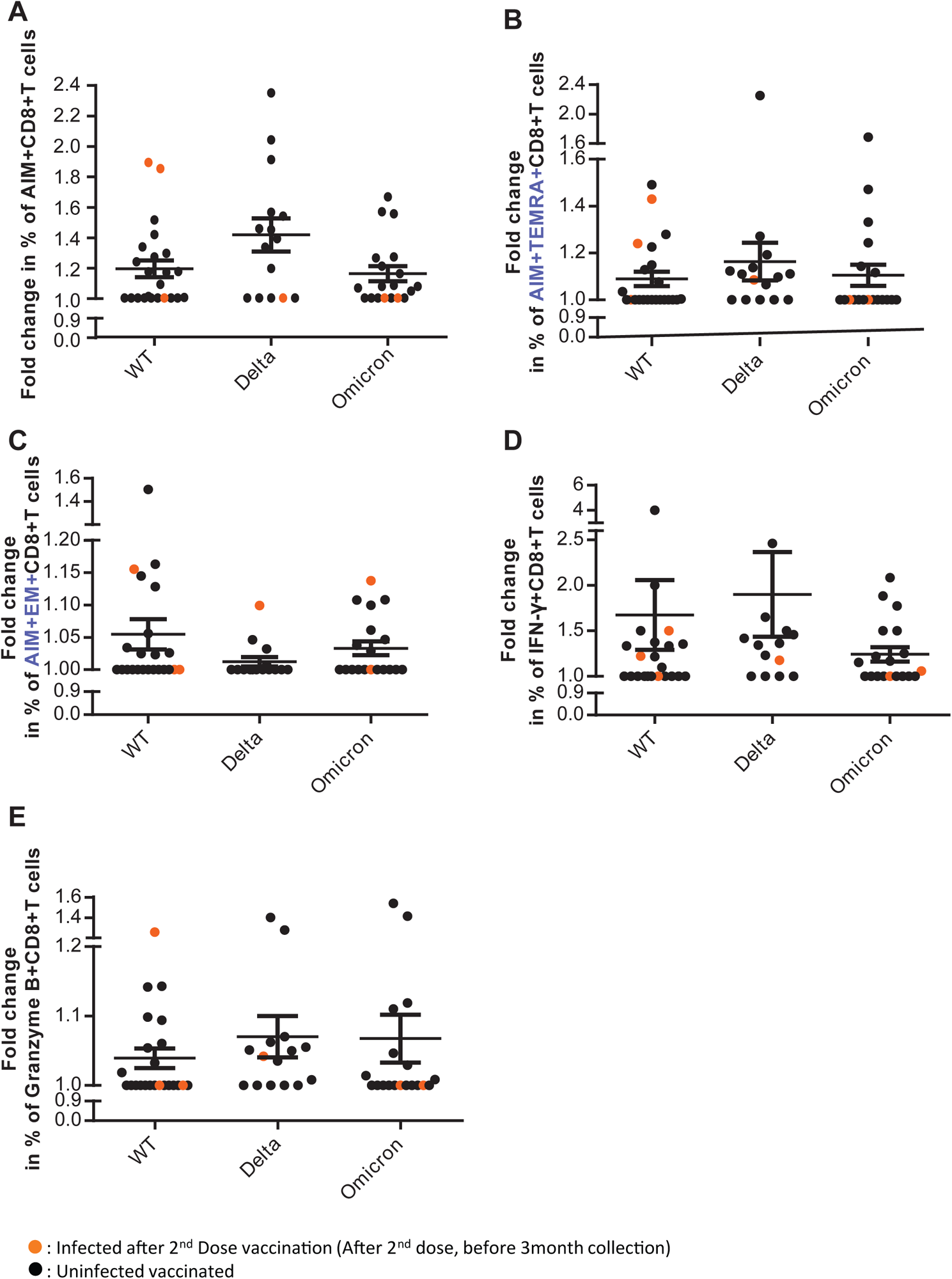
SARS-CoV-2 mutations and their activation competency in cytotoxic T cells following 3^rd^ dose vaccination. The graphs represented the effect of multiple SARS-CoV-2 variants upon the immunological responses for (A) AIM+, (B) AIM+TEMRA+, (C) AIM+EM+, (D) IFN-γ+ and (E) Granzyme B+ in CD8+ T cells in individuals who were administered their 3^rd^dose of vaccine. Data represented as mean ± SEM. n= 23 for spike WT, n= 15 for delta, n= 19 for omicron.

### Covaxin administration generated optimal CD4+ and CD8+Tcell responses in 33% of vaccinated subjects

As per results presented above, it was clear that Covaxin induces mild to moderate immune responses in T-cells. To estimate the frequency of subjects that were optimally responding to the Covaxin, we calculated stimulation index (SI) for CD4+ and CD8 T+cells. The magnitude of SARS-CoV-2 reactive T cell responses was measured using SI taking AIM+ expression into consideration [15]. All of the subjects were recruited between January 2021 to January 2023, which included convalescent as well as uninfected vaccinees. The stimulation index for CD4+ T cells and CD8+ T cells were calculated with limit of stimulation (LOS) of 1.84 and 1.27 for CD4+T (Figure 6 A) and CD8+ T cells (Figure 6 B) respectively. Samples with a value more than the cut off were assigned as responders and below the LOS were the low-responders. The analysis revealed that only 33% of the individuals in the total population (n=121) were optimal responders upon Covaxin administration.

**Figure 6:**
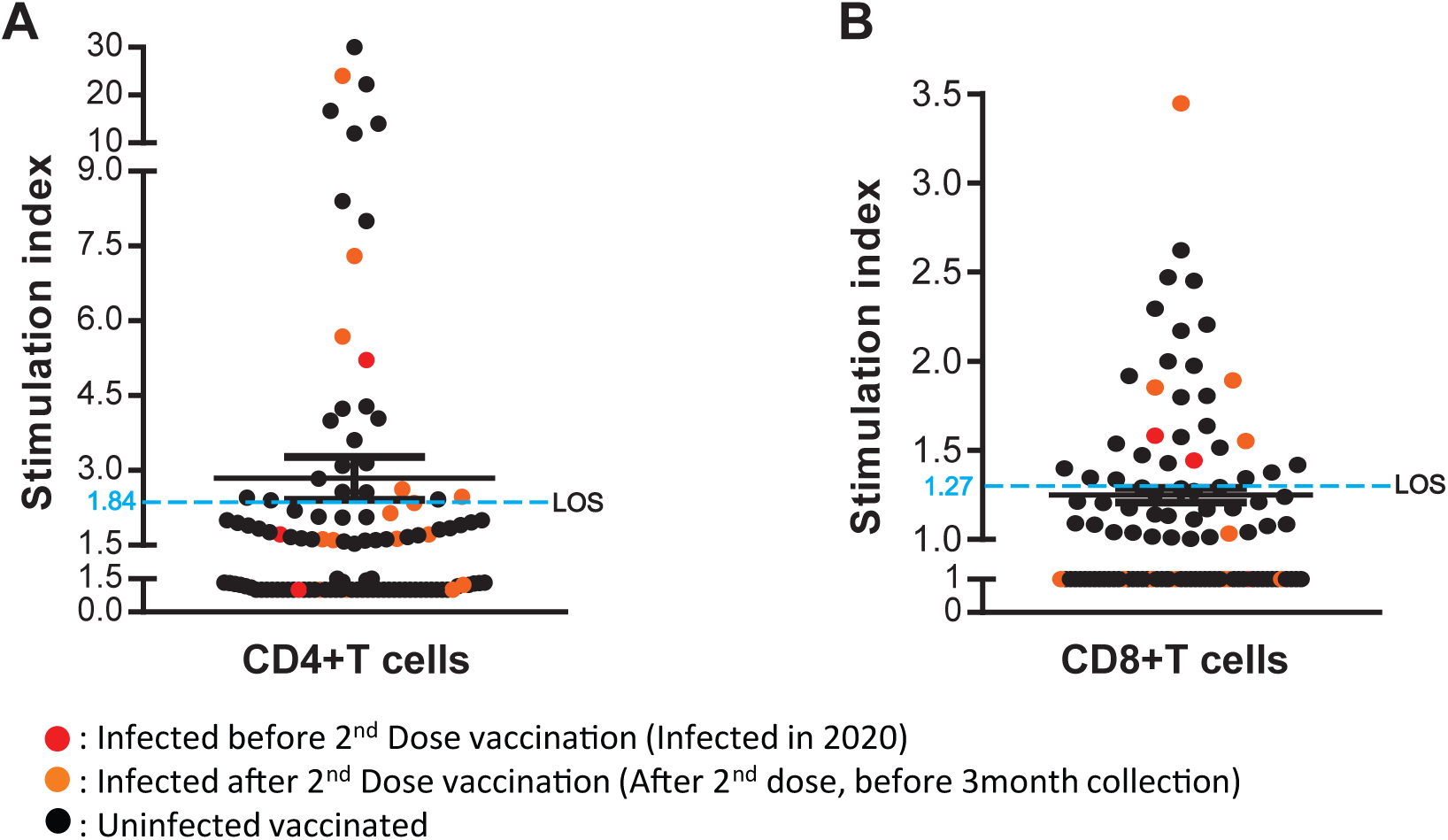
Stimulation index for CD4+ T cells and CD8+ T cells. The stimulation indices for (A) CD4+T and (B) CD8+ T cells have been expressed after calculation of the cut off values and setting the LOS (n=121). Individual data points are shown here as a scatter dot plot with lines showing the LOS. Data are expressed as geometric mean± SEM.

## Discussion

The current study attempted to bridge a substantial gap regarding the understanding of the longitudinal impact of emerging SARS-CoV-2 infection or inactivated virus-based vaccine administration on humoral and cell-mediated immunity. Here, we concentrated on analysing antibody and T-cell responses in a cohort of Covaxin vaccinated subjects as a follow-up study for 2 years. Full spectra multicolor flow cytometry was used to evaluate the CD4+ and CD8 +T cell responses and their subtypes and effector function in response to Covaxin and its cross-reactivity with Delta and Omicron SARS-CoV-2 variants. In addition, we estimated the titre of IgG antibody in a longitudinal manner and the virus neutralization capacity of these serum samples. To the best of our knowledge, this study reports for the first time about the longitudinal 2 years follow-up profiling of spike RBD antibody and T-cells responses after administration of an inactivated virus-based vaccine Covaxin (BBV152).

According to our longitudinal sero survey’s findings, the IgG antibody titer levels were significantly increased after 2^nd^ dose of Covaxin administration. After the 1^st^ dose, nearly 40-45% of subjects were non-responders. Overall, 15-20% of Covaxin vaccinees did not respond at all after vaccination. We also observed a gradual decrease in IgG titre after vaccination which was boosted strongly upon Omicron infection wave in January 2022. Omicron infection induced strong antibody response in almost 100% subjects and the antibody titres were maintained till one year. This finding suggested that the increase in seroprevalence among a large proportion of the population was due to exposure and natural infection with the Omicron variant during the third wave of COVID-19 in India that prevailed between January and March 2022. During the third wave of COVID-19, more than 20 million COVID-19 cases were reported from India, with the Omicron variant being a predominant circulating variant of concern [16–19]. These results, which were seen in convalescent and uninfected vaccine recipients, were undoubtedly caused by both a natural infection as well as COVID-19 vaccination [19–21]. After getting a sharp increase in antibody titre at 12 months after second dose and Omicron infection, our aim was to examine the neutralizing response at two-time intervals, namely before existence of delta variant (3 months) and after omicron infection (12 months) (Figure 1D). We found that 99% of study participants had a complete virus neutralising response after omicron infection, but more research is needed to determine whether neutralisation titres correlate with receptor-binding domain IgG antibodies.

The fundamental understanding of T cell responses to SARS-CoV-2 at various stages of immunisation is crucial. Here, we used PBMCs collected from vaccinated subjects to provide functional validation of CD4+ and CD8+T cell responses. Spike-specific peptide pools of predominant SARS-CoV-2 variants were included in the investigations.

We demonstrated that the CD4+T cells predominantly contribute to the cross reactivity against SARS-CoV-2 and the memory responses, with a little contribution from CD8+T cells (Figure 2 and 4). Other cohorts also showed the limited contribution of CD8+T cells among the cross-reactive T cells [22, 23]. The majority of Covaxin vaccinated and/or convalescent individuals who recovered from moderate disease lack SARS-CoV-2 specific CD8+T cells responses, which may be a result of poor memory CD8+T cell formation or poor stability. Understanding the role of CD8+T cells in the pathogenesis of SARS-CoV-2 will require further research into various illness outcomes across diverse populations. The antigen-specific T cell response was pre dominated by a Th1 phenotype with a substantial rise in cytokines, such as TNF-α and IFN-γ (Figure 2). In agreement with this, earlier research revealed a minimal Th2 response in COVID-19-positive, vaccinated people, confirming our findings [24–27]. The change in effector memory response (Figure 2 B) was determined across all five time points, however, insignificant and variable response was observed. Importantly, majority of the participants showed memory phenotype of virus-specific CD4+T cells in detectable limits persisted for at least up to 6 months aligns with previous report who had received BBV152 vaccine [27–29].

The collection of samples for this study (Figure 3), was performed at a time when the delta and Omicron VoCs existed, this allowed us to ascertain how vaccine induced CD4+T cells react with the spike antigen of ancestral strain, delta and omicron variants. Our data indicated that the extent of the antigen-specific T cell response against all variants in our investigation was comparable, despite multiple alterations in the delta and omicron variants [30, 31]. This is probably due to the earlier experienced T lymphocytes with conserved epitope region of ancestral strain, which helps in cross reacting with emerging variants [32, 33]. Furthermore, we observed that memory as well as Th1 response elicited during the active ancestral SARS-CoV-2 infection, but is relatively reduced against the delta and omicron variants, which was similar to earlier reports [34–36]. In contrast to earlier results in vaccinated people, our cohort (Figure 5) shows a more pronounced reduction in the magnitude of the CD8+T cells response to develop VoCs [32]. This finding might be explained by the fact that, the overall strength of CD8+T cell response in vaccinated samples is weak, which may have limited potential for cross-reactivity with the VoCs.

Participants who received the vaccine consistently produced a significant CD4+T cell response against SARS-CoV-2. Using stimulation index as the criterion, similar outcomes were reached (Figure 6). The analysis revealed that only ∼33% of the individuals in the total population were responders which was less than the published report’s finding that 50% of donors were beyond the limit of detection [22, 23]. This may have occurred because the sample size was constrained for practical reasons or vaccination is not enough to generate active immunization.

The next generation of vaccines must pay close attention to the extent of T cell responses induced upon vaccination given the significance of T-cell memory in limiting disease severity and safeguarding against emerging variants. However, few studies [33, 37] have looked into the factors contributing to strong T cell immune responses against SARS-CoV-2. When this study was planned, limited information was available about the vaccine induced cell mediated immune response. Since the main goal of the study was to track T cell’s protective immunity against this virus for a number of months after immunisation.

The COVID-19 incidence burden on the Indian sub-continent is significant, despite the fact that case fatality rates are very low: this could be due to the presence of highly cross reactive CD4+T cells. Understanding the role of cross-reactive CD4+T cells in disease outcome and immunological memory formation is crucial for COVID-19 vaccine development and implementation.

### Author contributions

Conceptualization, AT, SP, and SKR; Investigation, AT, SP, SS, and SKR; Analysis, AT, and SP; Sample management, SS, and SP; Writing, AT, SS and SKR.; Supervision and Funding, SKR. All study data were completely accessible to all authors. The submitted version of the article was reviewed and approved by all the authors.

## Data Availability

All data produced in the present study are available upon reasonable request to the authors

## Acknowledgments

We are thankful to all the participants for generous support in this study. For the technical assistance, we thank Ms. Rasmita Das, Ms. Suravi Mohanty, Mr. Tejeswar Dass, Mr. Dushmanta Parida and Sudarshana Jena (ILS, Bhubaneswar). We would also like to thank RMRC, Bhubaneswar for providing CLIA facility support for IgG antibody titre profiling. We are grateful to Department of Biotechnology, Biotechnology Industry Research Assistance Council (BT/CS0053/05/21), and ILS core grant for providing the financial support ensuring the smooth progress of the project.

