## Supplementary Info for "A prospective two-year longitudinal follow-up study depicting humoral and cell-mediated immune responses in Covaxin vaccinated individuals"

**Supplementary figure 1**

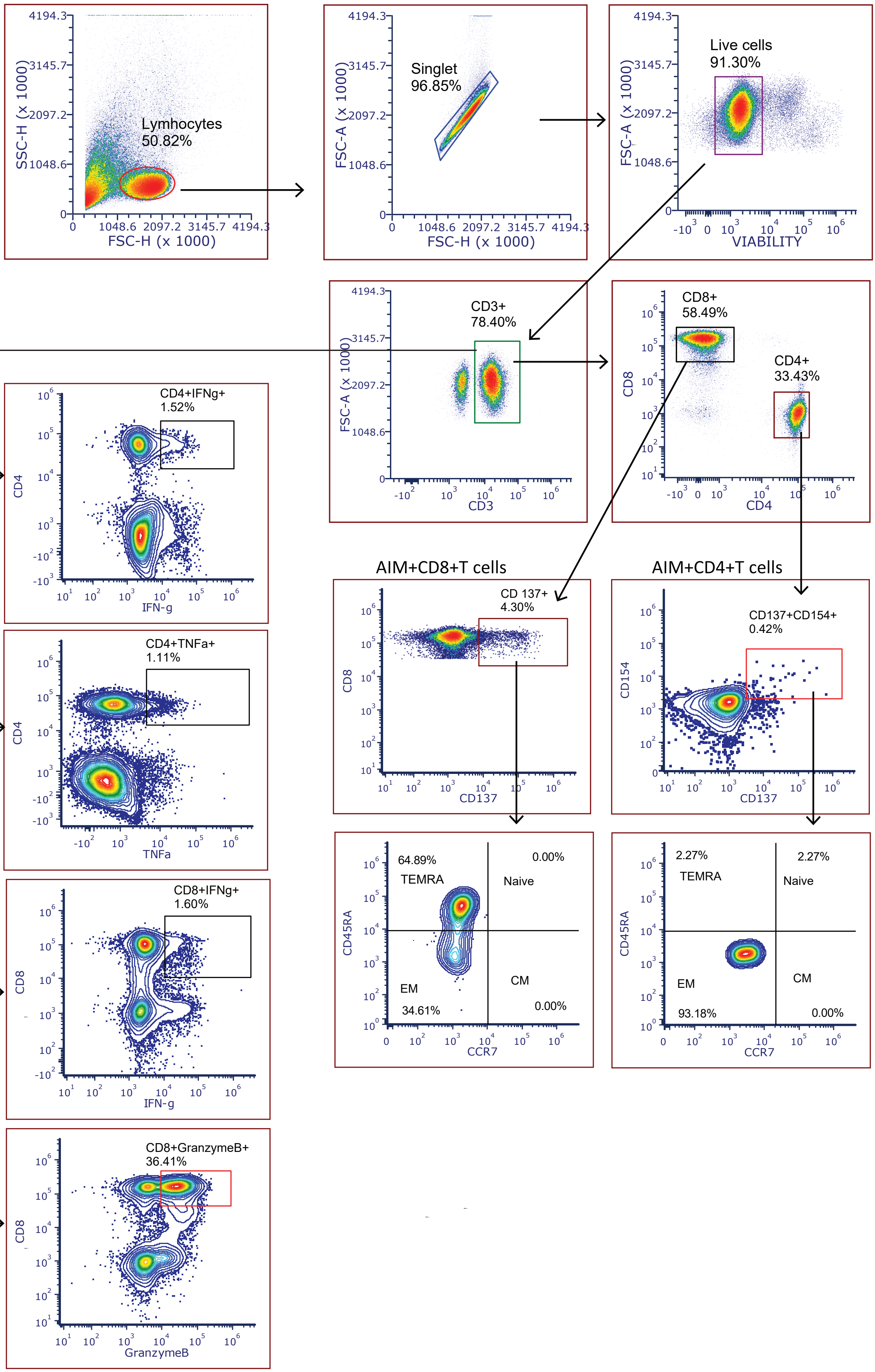

Supplementary Figure 1: Gating criteria used for determination of antigen-specific CD4+T and CD8+T cells, antigen-specific memory CD4 and CD8+ T-cell subsets, and cytokine profiling. Cells were differentiated into antigen-specific CD4+ T cells determined by CD137+CD154+(AIM+) and CD8+ T cells determined by CD137+ cells in the total CD4+T cells and CD8+T cells. Cells were differentiated into central memory (CM: CD45RACCR7+), effector memory (EM: CD45RACCR7+), naive (CD45RA+CCR7+), or terminally differentiated effector memory cells re-expressing CD45RA (TEMRA: CD45RA+CCR7−) out of AIM+ cells. IFNY, TNFα, Granzyme B expressing CD4+ and CD8+T cells were gated from CD3+ population.

#### Supplementary figure 2

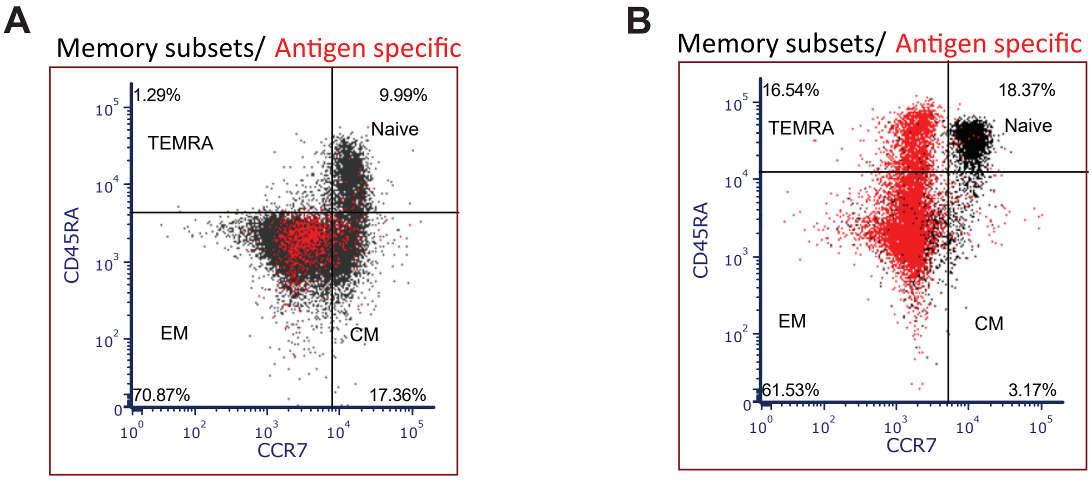


Supplementary Figure 2 : Representative FACS plots showing the AIM + cells (red) over total memory subsets (grey), determined as central memory (CM: CD45RA−CCR7+), effector memory (EM: CD45RA−CCR7−), naive (CD45RA+CCR7+) or terminally differentiated effector memory cells re-expressing CD45RA (TEMRA: CD45RA+CCR7−) in CD4+T (Figure A) cells and CD8+T (Figure B) after stimulation with peptide pool.

### Supplementary figure 3

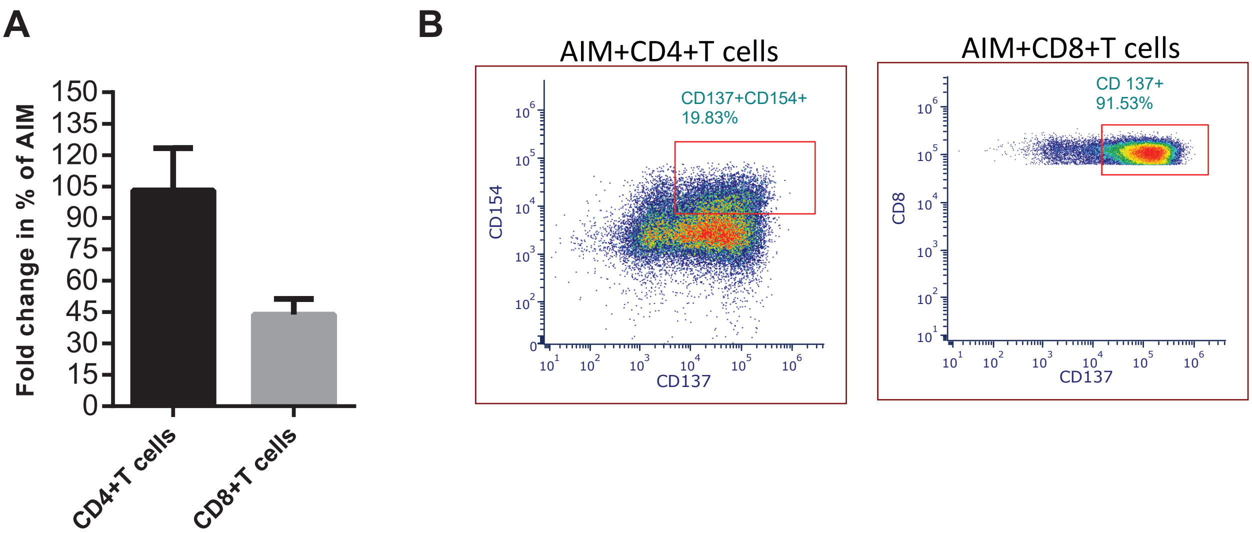


Supplementary Figure 3 : Bar plot represents the increase in fold change in % of AIM+CD4+ and CD8+T cells after stimulation with Cytostim (Figure A). Representative flow images also justify the same (Figure B).

#### Supplementary figure 4

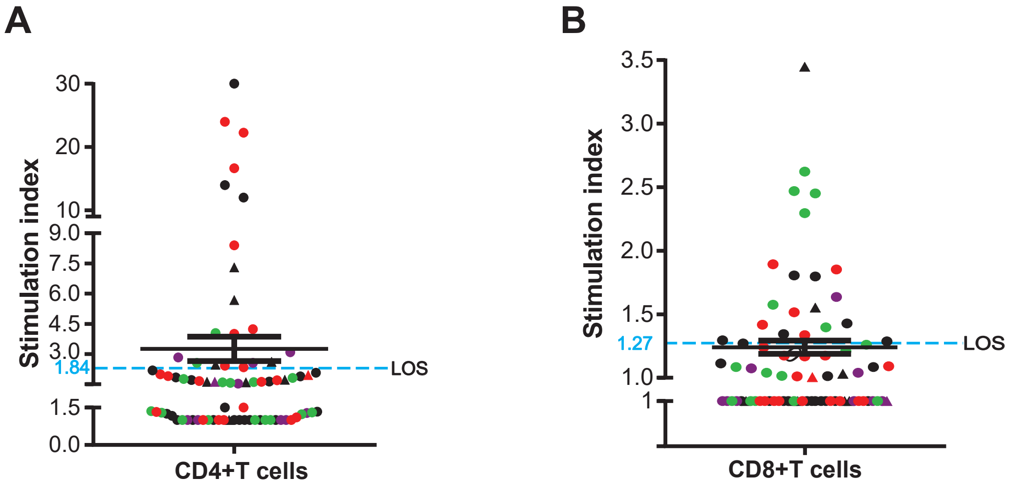


Supplementary Figure 4 : The Limit of Stimulation Index (LOS) was used to distinguish between responders and nonresponders and recovered vaccine recipients are denoted by triangles when the majority of them are below the cutoff line in case of both CD4+(Figure A) and CD8+ T cells (Figure B).

#### Supplementary figure 5

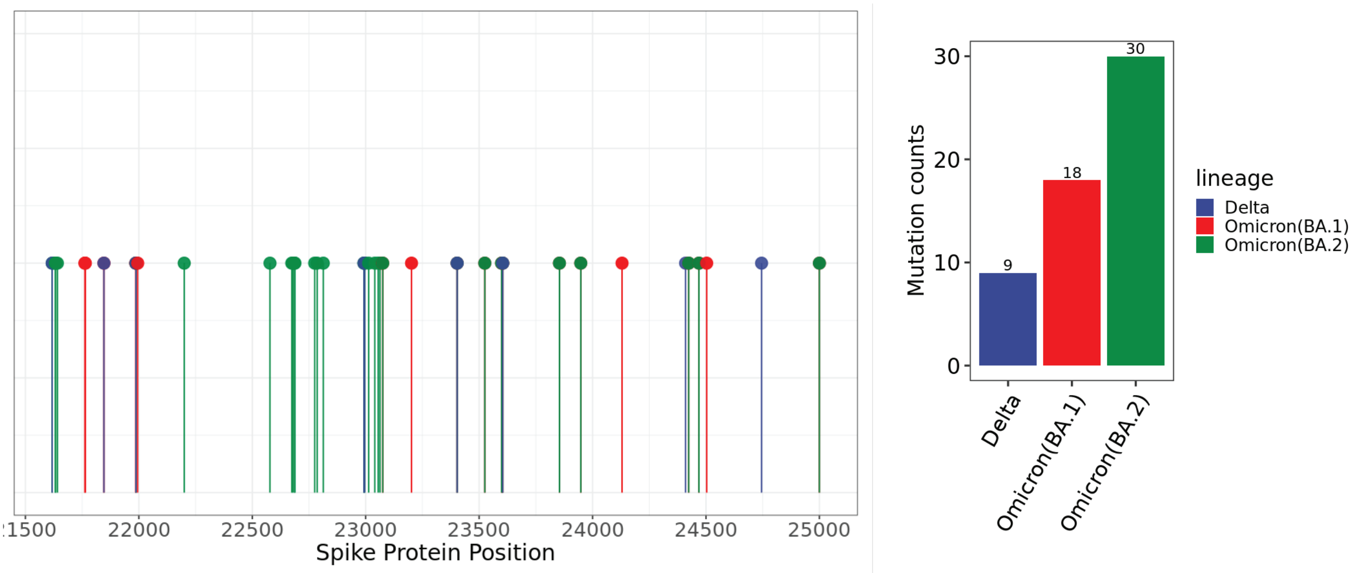


Supplementary Figure 5 : Mutation load distribution and mutation frequency in VoCs of SARS-CoV-2.
